# A cluster-randomized trial of client and provider-directed financial interventions to align incentives with appropriate case management in retail medicine outlets: results of the TESTsmART Trial in western Kenya

**DOI:** 10.1101/2023.09.14.23295586

**Authors:** J. Laktabai, E. Kimachas, J. Kipkoech, D. Menya, D. Arthur, Y. Zhou, T. Chepkwony, L. Abel, E. Robie, M. Amunga, G. Ambani, M. Woldeghebriel, E. Garber, Nwamaka Eze, Pamela Mudabai, J.A. Gallis, Chizoba Fashanu, I. Saran, A. Woolsey, T. Visser, E.L. Turner, W. Prudhomme O’Meara

## Abstract

ACTs are responsible for a substantial proportion of the global reduction in malaria mortality over the last ten years. These reductions would not have been possible without publicly-funded subsidies making these drugs accessible and affordable in the private sector. However, inexpensive ACTs available in retail outlets have contributed substantially to their overconsumption. We test an innovative, scalable, and sustainable strategy to target ACT subsidies to clients with a confirmatory diagnosis. We supportead point-of-care malaria testing (mRDTs) in 39 retail medicine outlets in western Kenya and randomized them to three study arms; control arm offering subsidized RDT testing for 0.4USD, client-directed intervention where all clients who received a positive RDT at the outlet were eligible for a free (fully subsidized) first-line ACT, and a combined client and provider directed intervention where clients with a positive RDT were eligible for free ACT and outlets received 0.1USD for every RDT performed. Our primary outcome was the proportion of ACT dispensed to individuals with a positive diagnostic test. Secondary outcomes included proportion of clients tested at the outlet and adherence to diagnostic test results. 43% of clients chose to test at the outlet. Test results informed treatment decisions and resulted in targeting of ACTs to confirmed malaria cases – 25.3% of test-negative clients purchased an ACT compared to 75% of untested clients. Client-directed and client+provider-directed interventions did not offer further improvements, compared to the control arm, in testing rates (RD=0.09, 95%CI:-0.08,0.26) or dispensing of ACTs to test-positive clients (RD=0.01,95% CI:-0.14, 0.16). Clients were often unaware of the price they paid for the ACT leading to uncertainty in whether the ACT subsidy was passed on to the client. We conclude that mRDTs could reduce ACT overconsumption in the private retail sector, but incentive structures are difficult to scale and their value to private providers is uncertain.

## Introduction

Each year, approximately one third of the more than 247 million cases of malaria world-wide^1^, and hundreds of millions of cases of non-malaria febrile illness, seek care at chemists, pharmacies and drug shops, often referred to collectively as the private retail health sector. In most malaria-endemic countries, first-line artemisinin combination therapies (ACTs) for malaria treatment are widely available over-the-counter (OTC) in the retail health sector. When first introduced in the 2000s, prices for ACTs were very high, requiring subsidies to make them accessible to most malaria patients.^2^ Since then, unsubsidized ACT prices have declined, making OTC ACTs affordable even without public subsidies.^3, 4^ Declining prices of ACTs, whether subsidized or not, create a trade-off between access and targeting; lower prices improve uptake of effective therapies by those with malaria but also increase inappropriate use by those without malaria. Inappropriate use neglects the true cause of illness potentially to the detriment of the client, wastes subsidy dollars on clients who don’t need antimalarials, and increases the drug pressure that spurs the spread of antimalarial resistance. Curbing inappropriate use and targeting ACTs to malaria cases requires parasitological diagnosis which is virtually absent in the retail sector in most endemic countries ^5^. As a result, although nearly 25% of global ACT demand is from the private *retail* sector^6^, virtually all ACTs dispensed OTC are to clients without a parasitological diagnostic test. In addition, there is evidence to suggest that between 65-91% of ACTs^7–9^ dispensed over the counter are purchased by people without malaria.

Several studies have explored the potential role of malaria rapid diagnostic tests (mRDTs) in improving case management in the retail sector, with mixed results^10, 11^. In such studies, which mostly included subsidies for the mRDT and ACT, between 7-100% of suspected malaria cases chose to be tested with an mRDT. Uptake of testing was found to be very sensitive to both the price of the mRDT itself as well as the price of the ACT. Higher testing rates were observed when the price of the drug was very high relative to the price of the test. However, as many as 40% of tested clients purchased an ACT following a *negative* test result while up to 70% of clients who tested *positive* do not purchase an ACT. These studies show that fixed low prices of the ACT and the mRDT are not sufficient to ensure adequate testing uptake or adherence to the test result.

Previous studies in retail outlets share two important features – 1) ACTs were heavily subsidized for all customers and 2) there was no relationship between the mRDT result and the ACT subsidy. In our study, we aimed to link the mRDT and ACT subsidies through the use of a diagnosis-dependent ACT subsidy. We hypothesized that the use of price differentials could attract the provider and the client toward the right diagnostic and treatment course. First, the cost of the mRDT was set below that of the first-line ACT which makes the test an affordable option to determine the need for a more expensive drug. Second, we also used a conditional ACT subsidy whereby clients received a fully subsidized (free) ACT conditional on a positive malaria diagnostic test result. In the context of a conditional ACT subsidy, information from an mRDT becomes more valuable; a negative test would alleviate the need for an (unsubsidized) ACT and a positive test would ensure they receive an effective ACT for free. Finally, to help offset the investment of the provider’s time and the possibility of losing an ACT sale to a test-negative client, we also assessed the effect of a small payment to the outlet for each test performed.

Here we report the results of the Malaria Diagnostic Testing and Conditional Subsidies to Target ACTs in the Retail Sector trial in Kenya (TESTsmART Kenya), a three-arm cluster randomized trial designed to test the effect of conditional ACT subsidies and modest provider incentives on appropriate use of ACT in the retail sector in western Kenya.

## Methods

### Overall study design

The TESTsmART-Kenya trial was a three-arm cluster randomized trial carried out in two counties in western Kenya. Forty retail medicine outlets (clusters) were enrolled and randomized to one of three arms: a control arm, a client-directed conditional subsidy arm (CD), and an arm with a combined conditional subsidy plus provider incentive (CDPD) arm. Full details of the study design were previously reported in the study protocol^12^ and accompanying amendment by Woolsey et al^13^ and are summarized here. Results from the TESTsmART study in Nigeria (TESTsmART Nigeria) will be reported separately.

### Study area

The study was conducted in Bungoma and Trans Nzoia counties in western Kenya, where malaria transmission is perennial with seasonal peaks. Previously, we documented in this region that 70.6% of all recent fevers in the community reported taking artemether lumefantrine (AL, the first-line ACT in Kenya), for their illness, and 58.6% of these obtained their AL from a retail outlet^14^. Despite the very large volume of ACTs being obtained in the private retail sector, these outlets are not permitted to conduct mRDTs in Kenya ^15^.

### Intervention

The three intervention arms considered for evaluation are as follows:

1. Control arm: mRDTs were distributed through the study at a wholesale price to the retail outlet of 25 Kenyan Shillings (KES) or 0.25 USD per test, and mRDTs were offered to clients at a recommended retail price of KES 40 (0.40 USD) per test. The retail outlet owner/attendant is trained to use a mobile reporting app which was designed for the study to capture and upload an image of RDT results together with some patient characteristics. The app was downloaded on a mobile phone offered to all participating outlets. Each outlet received 500 KES (5.00 USD) of mobile data per month to facilitate data and image submission.
2. Client-directed (CD) intervention arm: in addition to the interventions implemented in the control outlets, clients visiting retail outlets in this arm who purchase an mRDT and receive a positive test result were eligible to receive a free ACT which was to be provided by the outlet. The outlet would be reimbursed by the study (conditional subsidy; cost equivalent to 150 KES or 1.50 USD for adults and 60 KES or 0.60 USD for children)
3. Combined client-directed + provider-directed (CDPD) intervention arm: in addition to the interventions implemented in the control outlets and CD intervention arm, the retail outlet owner received a small incentive (10 KES, approximately 0.10 USD) for each mRDT they conduct and report using the mobile app).

### Study outcome measures

The primary outcome was ACT consumption by parasitologically confirmed malaria cases, defined as the proportion of ACTs that were dispensed to malaria test-positive clients, including those who were tested at the outlet and those who came with documentation of a positive test result (either microscopy or mRDT) from another provider.

There were four secondary outcomes, all binary:

1. Use of malaria rapid diagnostic test: Proportion of suspected malaria cases that received an mRDT at the outlet, where a suspected case was any client who was tested with an mRDT or was untested but purchased an antimalarial (AM) of any type.
2. Adherence to mRDT result: Proportion of clients that were tested for malaria with an mRDT at the outlet whose treatment adhered to the mRDT result (tested positive with an mRDT and purchased ACT or tested negative with an mRDT and did not purchase AM)
3. Appropriate case management: Proportion of all suspected malaria cases (defined as in 1 above) that were for malaria with an mRDT at the outlet and adhered to the test results.
4. ACT use among the untested: Proportion of untested clients who purchased ACT.

The primary and secondary outcomes were collected during exit interviews of a random sample of eligible clients leaving retail outlets (see *Data collection* below). Table S1 summarizes the numerator and denominator for each outcome measure.

### Power calculations

The study was designed to attain a minimum of 80% power at an overall 5% significance level for each of two pre-specified comparisons between arms for the primary outcome: the combined intervention (CDPD) arm vs. the client-directed (CD) arm and the CDPD arm vs. the control arm. All power calculations were based on a formula from Moulton and Hayes for comparing two proportions under a cluster-randomized trial design with details previously published^12, 13^. Using the conservative Bonferroni correction for our two pre-specified comparisons and using pre-randomization data on malaria testing and treatment behavior reported in the app, we expected to achieve 80.2% power for our comparison of the CDPD arm vs. the CD arm, and greater than 95% power for our comparison of the CDPD arm vs. the control arm.

### Randomization and recruitment

Outlets were mapped, surveyed and enrolled as described in Woolsey et al^12, 13^. Following enrollment, retail outlets were randomized to arms by the study statistician. We started with a list of 40 participating outlets which were randomized to 3 arms. Given 40 outlets cannot be equally allocated to 3 arms, the extra outlet was assigned to the control arm. To avoid potential contamination of treatment effects, randomization was constrained such that any outlets in close proximity were assigned to the same arm. We defined this proximity based on being able to see one outlet when standing at the other, or about 0.5km. In practice, only two pairs of outlets met this criteria, one pair in the control arm and one pair in the CDPD arm.

The final number of outlets that completed the study was 39. Two outlets exited after initiation of the intervention, both from the CD arm - one exited after declining to permit exit interviews, and one relocated outside of the study area. One outlet was moved from the control arm to the CD arm and a new outlet was recruited into the CD arm giving 39 outlets equally distributed across the arms. All changes were made within the first five months of data collection and data from the exited outlets is not included in the analysis. Only data from the period following the change of arms is included for the outlet which was moved from control to CD arm.

### Study implementation

Enrolled outlets were trained on malaria case management guidelines, correct mRDT procedures, and the use of the study mobile app to record client encounter data and take photos of mRDT results. During a brief pilot period, the outlets started collecting data on the mobile app and performing mRDTs. Outlets were monitored closely to make sure testing and app issues were resolved before initiating the intervention. After the three month pilot period, outlets were trained in arm-specific groups on the intervention guidelines for conditional subsidies and provider-directed incentives, as applicable. They were also informed of the exit interview process being conducted outside of their outlet, independently of the client-outlet interaction. Outlets were provided with a small monthly stipend with which they were encouraged to engage a laboratory technician to perform RDTs at the outlet. The intervention continued for 18 months to allow for adequate burn in with the intention to reach a ‘steady state’.

The study team conducted regular supervision visits over the first six months to deliver mRDT kits, ensure good adherence to testing and safety procedures, troubleshoot challenges with the mobile app and provide onsite mentorship to all outlets. Thereafter, outlets ordered replenishment of mRDT kits as needed with deliveries on one scheduled day each week. Supervision and mentorship were conducted if outlets raised concerns or if there were any abnormal mRDT photographs observed in the encounters submitted via the app.

Subsidies and incentives were paid weekly to mobile money accounts of the owners of the intervention outlets. Outlets allocated to the CD or CDPD arms agreed to give free AL to test-positive clients for which they were reimbursed by the study (60 KES or 0.6US for children 9 years and below and 150 KES or 1.5USD for clients 10 years and above). Outlets allocated to the CDPD arm received a small payment for administering RDTs to clients (10 KES, approximately 0.10USD). Payments were calculated based on photographs of RDTs uploaded to the study app. Outlets were paid for total tests conducted (CDPD arm only) and for positive RDTs when the outlet also reported that AL was dispensed (both CD and CDPD arms). For this study, reimbursable AL was any brand of artemether lumefantrine in a tablet formulation (not syrup). Outlets were trained how to identify the drug ingredients and most common brands of AL.

During the intervention period, we observed that testing rates were lower in our ongoing study than in the pilot study ^16^. To ensure that clients were aware of the opportunity for testing and the conditional subsidy (if applicable), we deployed a client-facing sensitization at approximately the midpoint of the intervention period (October 2021, after 10 months of intervention monitoring). For the remaining 9 months, we enlisted local community health workers (CHWs) to distribute fliers to all clients entering a study outlet on randomly selected days, regardless of why they were visiting the outlet. The message on the flier was tailored to each arm and CHWs were instructed to keep their interaction with entering clients very brief and refer them to the outlet attendant if there were questions. The goal was to ensure clients were aware of the availability of testing and, in the intervention arms, the opportunity for a free AL conditional on a positive test.

### Data collection

Interviews were conducted between January 1, 2021 and August 31, 2022 with clients departing from participating retail outlets on random days of the week. Interviewers were blinded to arm assignment. All clients exiting the outlet that day were eligible to be screened. Clients were invited to participate in the exit interview if the patient experiencing symptoms was present at the outlet and older than 1 year; they were seeking treatment for malaria-like symptoms but did not have any symptoms of severe malaria; and they had not already taken an antimalarial in the last 7 days. For patients under the age of 18, an accompanying parent or guardian was asked to participate as the responding client. Data for participant exit interviews was collected electronically via tablet. The primary tool for developing the data collection forms was REDCap hosted at Duke University^17, 18^.

### Statistical analysis

Statisticians were blinded to arm identity until analysis was complete. Baseline characteristics were summarized by study arm using means and standard deviations for continuous variables, and counts and percentages for categorical variables. We analyzed all client-level self-reported primary and secondary outcomes using the modified Poisson approach^19, 20^ with Kauermann-Carroll corrected robust standard errors^21, 22^, with log link to estimate risk ratios (RRs) and identity link to estimate risk differences (RDs). This approach was implemented within the generalized estimating equations (GEE)^23, 24^ framework to account for clustering of outcomes by outlet (or by pair of outlets, in the case of the pair of “close” shops randomized to the same arm), and utilizing an independence working correlation matrix to avoid bias in estimating intervention effects^25^. We intended to estimate correlation parameters using the matrix-adjusted estimating equation (MAEE) approach^26^, but since this approach did not converge, we reverted to standard GEE with method of moments estimation of correlation parameters.

All models included fixed effects for the intervention arm indicators, the stratification variable (county), and time (in months, including linear, quadratic, and cubic terms) to account for potential imbalanced recruitment over the 20-month study period (referred to as minimally adjusted models). We also present fully adjusted models which include the following variables which may be imbalanced by intervention arm (potential confounder variables): client gender, client age, level of schooling, and wealth index. In addition, we pre-specified several sensitivity analyses for the primary outcome. Specifically, 1) to restrict ACT to only AL which was subsidized in the study, 2) to evaluate outcomes before and after expansion of the list of ALs that qualified for the study subsidy, 3) to evaluate outcomes before and after client-facing sensitization.

Inference for intervention effects was based on the *t*-statistic with degrees of freedom given by *I* – *p*, where *I* was the number of clusters (retail outlets) and *p* was the number of parameters in the mean model (specifically of the cluster-level covariates, including the intercept, treatment indicator, county, and time variables). Given that we have two pre-specified comparisons for each outcome, namely CDPD vs. CD, and CDPD vs. control arm, for streamlining of reporting, all tables show the CDPD arm as the reference. All analyses were based on the intention-to-treat principle. Since we did not have longitudinal follow-up of clients, we did not need to account for missing data due to attrition of clients. Clients missing specific data elements were excluded from models requiring those variables as indicated in the tables. Overall, missing data was minimal and highest for the household asset questions where 5.7% of participants were missing adequate information to construct their wealth score.

### Ethics statement

The study was reviewed and approved by Duke University Institutional Review Board (Pro00104256) and Moi University Institutional Research and Ethics Committee (IREC/2019/304). The study is registered in ClinicalTrials.gov (NCT04428307). Verbal informed consent was obtained from all participants and logged in the REDCap data collection form.

## Results

### Analysis population

Between January 2021 and August 2022, data were collected from customers exiting participating retail outlets on 5,416 outlet-days of observation. Out of 11,783 individuals approached when exiting the retail outlets, 5,952 were eligible to participate (50.5%; Figure 1). Of those eligible, 5,696 consented (95.7%). Of those not eligible, 23% (n=1,335) had taken antimalarials in the last 7 days and 61% (n=3,578) were buying drugs for someone not present, usually a child at home, and therefore unavailable for testing. Other reasons for exclusion included the absence of malaria-like symptoms (11%), less than one year of age (5.0%), or no adult present to consent for a minor (2.0%).

**Fig 1:**
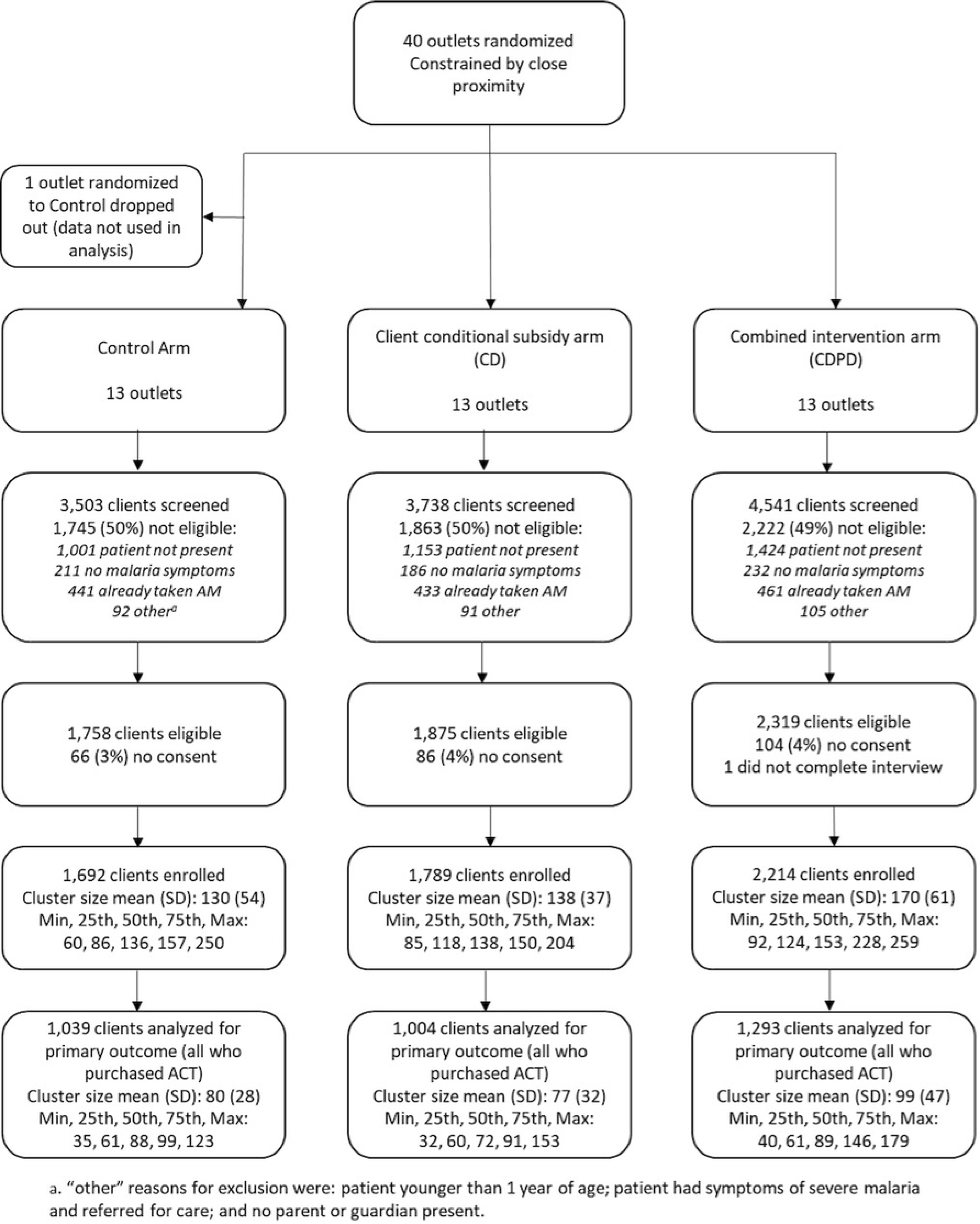
Flow diagram of shop enrollment, randomization and client interviews.

Of the 5,696 participants who were eligible, consented and completed the interview (one client discontinued the interview before any outcome data was collected), 69% of sick clients were adults and of those, most were between 40-59 years of age (Table 1). The proportion of male adult clients was slightly higher than female (54% vs. 46%) but respondents accompanying a child client were predominantly female (77%). 65% of adult clients and 65% of caretakers accompanying a minor had schooling beyond the primary level. Participant characteristics were similar across arms.

**Table 1:** Characteristics of participants enrolled in TESTsmART by arm.

|  | Study Arm |  |  |  |
| --- | --- | --- | --- | --- |
| Characteristic | Control | CD | CDPD | Overall |
| Interviewees, N<sup>1</sup> | 1,692 | 1,789 | 2,214 | 5,695 |
| Respondent, n (%) |  |  |  |  |
| Adult client | 1,151 (68%) | 1,292 (72%) | 1,502 (68%) | 3,945 (69%) |
| Guardian of child client | 541 (32%) | 497 (28%) | 712 (32%) | 1,750 (31%) |
| **Adult clients, n** | **1,151** | **1,292** | **1,502** | **3,945** |
| Age, n (%) |  |  |  |  |
| 18-25 | 202 (18%) | 195 (15%) | 216 (14%) | 613 (16%) |
| 26-39 | 377 (33%) | 447 (35%) | 506 (34%) | 1,330 (34%) |
| 40-59 | 424 (37%) | 490 (38%) | 589 (39%) | 1,503 (38%) |
| 60-79 | 133 (12%) | 143 (11%) | 177 (12%) | 453 (12%) |
| 80 and above | 12 (1.0%) | 15 (1.2%) | 10 (0.7%) | 37 (0.9%) |
| Missing | 3 | 2 | 4 | 9 |
| Gender, n (%) |  |  |  |  |
| Male | 617 (54%) | 708 (55%) | 792 (53%) | 2,117 (54%) |
| Female | 534 (46%) | 584 (45%) | 710 (47%) | 1,828 (46%) |
| Education level, n (%) |  |  |  |  |
| None | 61 (5.4%) | 56 (4.4%) | 65 (4.4%) | 182 (4.7%) |
| Primary | 314 (28%) | 427 (34%) | 424 (29%) | 1,165 (30%) |
| Secondary | 465 (41%) | 516 (41%) | 609 (41%) | 1,590 (41%) |

| Characteristic | Study Arm |  |  |  |
| --- | --- | --- | --- | --- |
|  | Control | CD | CDPD | Overall |
| College | 223 (20%) | 214 (17%) | 289 (20%) | 726 (19%) |
| University | 58 (5.2%) | 60 (4.7%) | 85 (5.8%) | 203 (5.3%) |
| Missing | 30 | 19 | 30 | 79 |
| **Child clients, n** | **541** | **497** | **712** | **1,750** |
| Respondent for child |  |  |  |  |
| Age, n (%) |  |  |  |  |
| 18-25 | 62 (11%) | 68 (14%) | 98 (14%) | 228 (13%) |
| 26-39 | 295 (55%) | 272 (55%) | 416 (59%) | 983 (56%) |
| 40-59 | 155 (29%) | 132 (27%) | 177 (25%) | 464 (27%) |
| 60-79 | 27 (5.0%) | 24 (4.8%) | 19 (2.7%) | 70 (4.0%) |
| 80 and above | 1 (0.2%) | 0 (0%) | 0 (0%) | 1 (<0.1%) |
| Missing | 1 | 1 | 2 | 4 |
| Gender, n (%) |  |  |  |  |
| Male | 136 (25%) | 112 (23%) | 156 (22%) | 404 (23%) |
| Female | 405 (75%) | 384 (77%) | 552 (78%) | 1,341 (77%) |
| Missing | 0 | 1 | 4 | 5 |
| Education level, n (%) |  |  |  |  |
| None | 19 (3.6%) | 17 (3.5%) | 18 (2.6%) | 54 (3.2%) |
| Primary | 171 (32%) | 165 (34%) | 216 (31%) | 552 (32%) |
| Secondary | 228 (43%) | 214 (44%) | 297 (43%) | 739 (43%) |
| College | 96 (18%) | 78 (16%) | 132 (19%) | 306 (18%) |
| University | 18 (3.4%) | 12 (2.5%) | 26 (3.8%) | 56 (3.3%) |
| Missing | 9 | 11 | 23 | 43 |
| Child client age |  |  |  |  |
| Mean (SD) | 7.0 (4.6) | 6.8 (4.9) | 6.3 (4.7) | 6.7 (4.7) |
| Missing | 1 | 2 | 0 | 3 |
| Child client gender, n (%) |  |  |  |  |
| Male | 279 (52%) | 259 (52%) | 361 (51%) | 899 (51%) |
| Female | 262 (48%) | 238 (48%) | 351 (49%) | 851 (49%) |
| **All clients, n** | **1,692** | **1,789** | **2,214** | **5,695** |
| Wealth index (quintile), n (%) |  |  |  |  |
| 0 to 20th | 342 (22%) | 349 (21%) | 387 (18%) | 1,078 (20%) |
| >20.0 to 40th | 320 (20%) | 365 (22%) | 388 (18%) | 1,073 (20%) |
| >40.0 to 60th | 316 (20%) | 324 (19%) | 440 (21%) | 1,080 (20%) |
| >60.0 to 80th | 352 (22%) | 313 (19%) | 470 (22%) | 1,135 (21%) |
| >80.0 | 260 (16%) | 329 (20%) | 415 (20%) | 1,004 (19%) |
| Missing | 102 | 109 | 114 | 325 |
| Time Period, n (%) |  |  |  |  |
| Jan-May 2021 | 294 (17%) | 276 (15%) | 367 (17%) | 937 (16%) |
| Jun-Oct 2021 | 572 (34%) | 619 (35%) | 710 (32%) | 1,901 (33%) |
| Nov 2021-Mar 2022 | 437 (26%) | 474 (26%) | 626 (28%) | 1,537 (27%) |
| Apr-Aug 2022 | 389 (23%) | 420 (23%) | 511 (23%) | 1,320 (23%) |
|  | Control | CD | CDPD | Overall |
| <sup>1</sup>Includes only interviewees who consented and met all inclusion criteria. One participant did not complete enough of the interview to contribute to any of the outcomes, and is not included in this table. |  |  |  |  |

### Malaria testing

Across all outlets, 61% (n=3474/5695) of clients who were interviewed had a malaria diagnostic test before purchasing drugs; 43% (n=2436/5695) were tested at the outlet, while 18.2% (n=1039/5695) were tested elsewhere before coming to the outlet. Prior to the study, testing was not offered in any of the retail outlets, but during the study period 52% (n=2378/4598) of those who did not have a prior diagnosis chose to be tested before purchasing treatment (Table 2). Testing rates were similar across wealth quintiles (Table S3b), however, the lowest wealth quintiles had lower testing proportions at the outlet (Table S2). Testing rates did not differ by gender of the child or by gender of the respondent accompanying a child, however, adult female clients had higher testing rates than their male counterparts (46% vs. 31%, Table S2). Higher testing rates were observed for children than for adults, particularly for school-age children (38% for adults, 56% for children 5-17 years; Table S2).

**Table 2:** Testing and treatment decisions by arm.

| Characteristic | Study Arm |  |  |  |
| --- | --- | --- | --- | --- |
|  | Control | CD | CDPD | Overall |
| Interviewees, n<sup>a</sup> | 1,692 | 1,789 | 2,214 | 5,695 |
| **Tested<sup>b</sup> at outlet, n** | 819 | 804 | 813 | 2,436 |
| *Proportion tested at outlet* | *(48%)* | *(45%)* | *(37%)* | *(43%)* |
| Positive test at outlet, n | 375 | 248 | 219 | 842 |
| *Proportion with positive test* | *(46%)* | *(31%)* | *(27%)* | *(35%)* |
| Took AL, n (%) | 185 (49%) | 150 (60%) | 116 (53%) | 451 (54%) |
| Took non-AL ACT, n (%) | 68 (18%) | 37 (15%) | 56 (26%) | 161 (19%) |
| Took other antimalarial, n (%) | 18 (4.8%) | 9 (3.6%) | 6 (2.7%) | 33 (3.9%) |
| Negative test at outlet, n | 428 | 545 | 581 | 1,554 |
| *Proportion with negative test* | *(52%)* | *(68%)* | *(71%)* | *(64%)* |
| Took AL, n (%) | 97 (23%) | 113 (21%) | 124 (21%) | 334 (21%) |
| Took non-AL ACT, n (%) | 17 (4.0%) | 13 (2.4%) | 29 (5.0%) | 59 (3.8%) |
| Took other antimalarial, n (%) | 9 (2.1%) | 11 (2.0%) | 9 (1.5%) | 29 (1.9%) |
| Invalid/unknown test result at outlet, n | 16 | 11 | 13 | 40 |
| **Not tested at outlet, n** | 873 | 985 | 1,401 | 3,259 |
| *Proportion not tested at outlet* | *(52%)* | *(55%)* | *(63%)* | *(57%)* |
| No test, n | 676 | 746 | 798 | 2,220 |
| Took AL, n (%) | 457 (68%) | 503 (67%) | 527 (66%) | 1,487 (67%) |
| Took non-AL ACT, n (%) | 66 (9.8%) | 39 (5.2%) | 74 (9.3%) | 179 (8.1%) |
| Took other antimalarial, n (%) | 50 (7.4%) | 80 (11%) | 74 (9.3%) | 204 (9.2%) |

<sup>b</sup> This number includes 58 interviewees who reported a test from elsewhere and also tested at the outlet. Test=mRDT

|  |  |  |  |  |
| --- | --- | --- | --- | --- |
| Had positive test from elsewhere, n | 154 | 156 | 403 | 713 |
| Took AL, n (%) | 71 (46%) | 87 (56%) | 186 (46%) | 344<sup>a</sup> (48%) |
| Took non-AL ACT, n (%) | 57 (37%) | 32 (21%) | 116 (29%) | 205<sup>b</sup> (29%) |
| Took other antimalarial, n (%) | 7 (4.5%) | 10 (6.4%) | 27 (6.7%) | 44 (6.2%) |
| Had negative test from elsewhere, n | 42 | 83 | 198 | 323 |
| Took AL, n (%) | 8 (19%) | 17 (20%) | 47 (24%) | 72 (22%) |
| Took non-AL ACT, n (%) | 4 (9.5%) | 4 (4.8%) | 9 (4.5%) | 17 (5.3%) |
| Took other antimalarial, n (%) | 1 (2.4%) | 0 (0%) | 3 (1.5%) | 4 (1.2%) |
| Unknown test result from elsewhere, n | 1 | 0 | 2 | 3 |

<sup>a</sup> This number includes interviewees who reported a positive test from elsewhere but did not provide documentation. The number who provided documentation of a positive test (required to be included in numerator of the primary outcome) and purchased AL was 181.
<sup>b</sup> This number includes interviewees who reported a positive test from elsewhere but did not provide documentation. The number who provided documentation of a positive test (required to be included in numerator of the primary outcome) and purchased ACT (non-AL) was 109.

Test positivity rate for the study period for tests conducted at the outlet as reported by the respondent was 35% (Table 2). Among clients with a prior test, 69% (n=713/(713+326)) reported a positive result.

### ACT consumption

Across the study arms, we observed that testing was associated with better targeting of ACTs (AL or other ACT; Table 2, Figure 2). Specifically, ACT consumption was markedly lower among those with a negative malaria test at the outlet (25.3%, n=393/1554) than among clients who did not test (75%, n=1666/2220). Those who reported a positive test at the outlet had similar ACT consumption to those without a test (73%, n=612/842). Although adult female clients were more likely to test, they did not have higher adherence to test results (Table S2).

**Fig 2:**
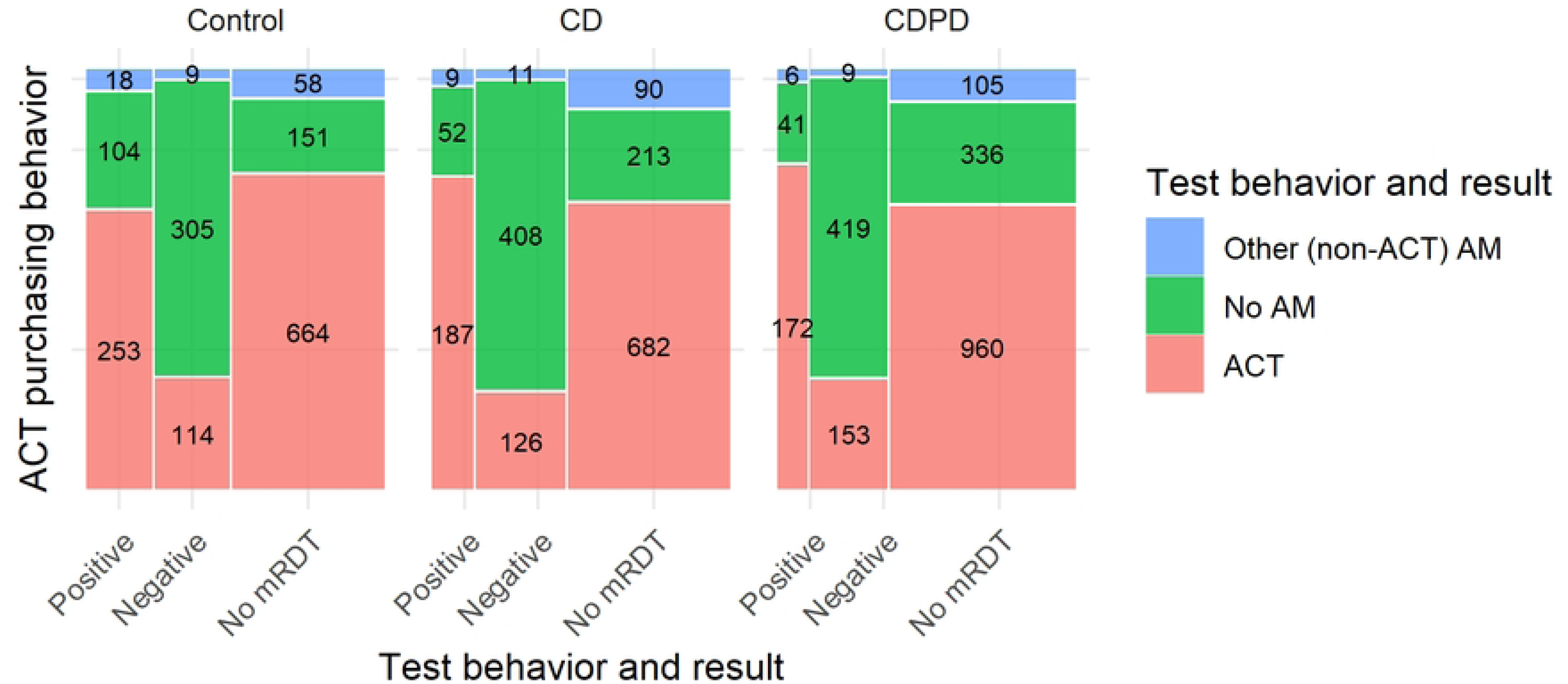
Malaria testing and antimalarial consumption by arm. ACT includes AL plus all other artemisinin-combination therapies.

### Intervention effects

Our primary outcome was chosen to measure the effect of the interventions on both improving testing uptake and targeting of ACTs which should result in a higher proportion of ACTs sold appropriately to test-positive clients. Overall, only 27% (n=902/3336) of all ACTs sold to participants was dispensed to malaria test-positive clients. We found no statistically significant impact of the conditional subsidy (CD) or the combined intervention (CDPD) on the primary outcome (Control: CDPD Adj. RR=1.11 [95%CI: 0.61-2.02], Table 3).

**Table 3:** Minimally adjusted and fully adjusted estimates of intervention effects for pre-specified study outcomes.

| Arm | Proportion | Risk Ratio (95% CI) |  | Risk Difference (95% CI) |  |
| --- | --- | --- | --- | --- | --- |
|  |  | Minimally Adjusted | Fully Adjusted | Minimally Adjusted | Fully Adjusted |
| Primary – ACTs sold to malaria test-positive clients |  |  |  |  |  |
| Control | 313/1,039<br>(30%) | 1.11<br>(0.61, 2.02) | 1.15<br>(0.71, 1.87) | 0.005<br>(-0.16, 0.17) | 0.01<br>(-0.14, 0.16) |
| CD | 250/1,004<br>(25%) | 0.92<br>(0.49, 1.71) | 0.94<br>(0.57, 1.55) | -0.05<br>(-0.22, 0.11) | -0.05<br>(-0.20, 0.09) |
| CDPD | 339/1,293<br>(26%) | — | — | — | — |
| Secondary 1 -- Suspected malaria cases that receive a mRDT (testing uptake) |  |  |  |  |  |
| Control | 819/1,541<br>(53%) | 1.22<br>(0.83, 1.80) | 1.20<br>(0.85, 1.70) | 0.09<br>(-0.11, 0.29) | 0.09<br>(-0.08, 0.26) |
| CD | 804/1,576<br>(51%) | 1.15<br>(0.81, 1.62) | 1.14<br>(0.83, 1.56) | 0.04<br>(-0.14, 0.21) | 0.07<br>(-0.09, 0.22) |
| CDPD | 813/1,878<br>(43%) | — | — | — | — |
| Secondary 2 – Malaria tested clients whose treatment adhered to test results |  |  |  |  |  |
| Control | 558/819<br>(68%) | 0.94<br>(0.80, 1.09) | 0.94<br>(0.81, 1.10) | -0.06<br>(-0.17, 0.05) | -0.05<br>(-0.16, 0.07) |
| CD | 595/804<br>(74%) | 0.99<br>(0.85, 1.16) | 1.00<br>(0.86, 1.17) | -0.01<br>(-0.13, 0.11) | 0.00<br>(-0.12, 0.11) |
| CDPD | 591/813<br>(73%) | — | — | — | — |
| Secondary 3 – Suspected malaria cases that are managed appropriately |  |  |  |  |  |
| Control | 558/1,541<br>(36%) | 1.14<br>(0.75, 1.73) | 1.13<br>(0.77, 1.66) | 0.03<br>(-0.12, 0.17) | 0.03<br>(-0.10, 0.16) |
| CD | 595/1,576<br>(38%) | 1.15<br>(0.77, 1.74) | 1.15<br>(0.78, 1.70) | 0.02<br>(-0.13, 0.17) | 0.04<br>(-0.10, 0.17) |
| CDPD | 591/1,878<br>(31%) | — | — | — | — |
| Secondary 4 – Untested clients taking ACT |  |  |  |  |  |
| Control | 523/676<br>(77%) | 1.03<br>(0.95, 1.11) | 1.01<br>(0.93, 1.10) | 0.02<br>(-0.04, 0.08) | 0.01<br>(-0.06, 0.07) |
| CD | 542/746<br>(73%) | 0.96<br>(0.89, 1.04) | 0.96<br>(0.89, 1.03) | -0.03<br>(-0.09, 0.03) | -0.03<br>(-0.08, 0.02) |
| CDPD | 601/798<br>(75%) | — | — | — | — |

As with the primary outcome, we did not observe any differences between arms for the secondary outcome of malaria testing. There was some evidence of higher testing in the control arm (53% compared to 43% in the CDPD intervention arm, Table 3), however, this did not reach statistical significance in the models (Control: CDPD Adj RR=1.22 [95%CI: 0.83-1.80]). Between 68-74% of those who tested adhered to the results of the test, with no significant differences between arms (Control: CDPD Adj RR=0.94 [95%CI: 0.80-1.09]). Similarly, there was no measurable effect of the intervention on other secondary outcomes of appropriate management and ACT use among untested clients.

Test positivity rates were substantially lower in the intervention arms (46% in the control arm compared to 27% in CDPD arm and 31% in CD, Table 2 and Figure 2). Lower test-positivity but higher ACT consumption by test-positives in the intervention arms (75% CD or 79% CDPD vs. 67% control, Table 2) resulted in only very small changes to the composite outcomes of appropriate management and adherence. These changes were not statistically distinguishable from the control arm.

### Understanding the intervention

In order to understand whether there was variability in the adoption of mRDTs at the study outlets, we examined differences in testing and treatment at the outlet (cluster) level. The outlets were very diverse with respect to their case management strategy (Figure 3). Testing rates ranged from as low as 10% of suspected cases to more than 80% with no discernable pattern by arm. Similarly, adherence to the test result ranged from 40 to 97% by outlet.

**Fig 3:**
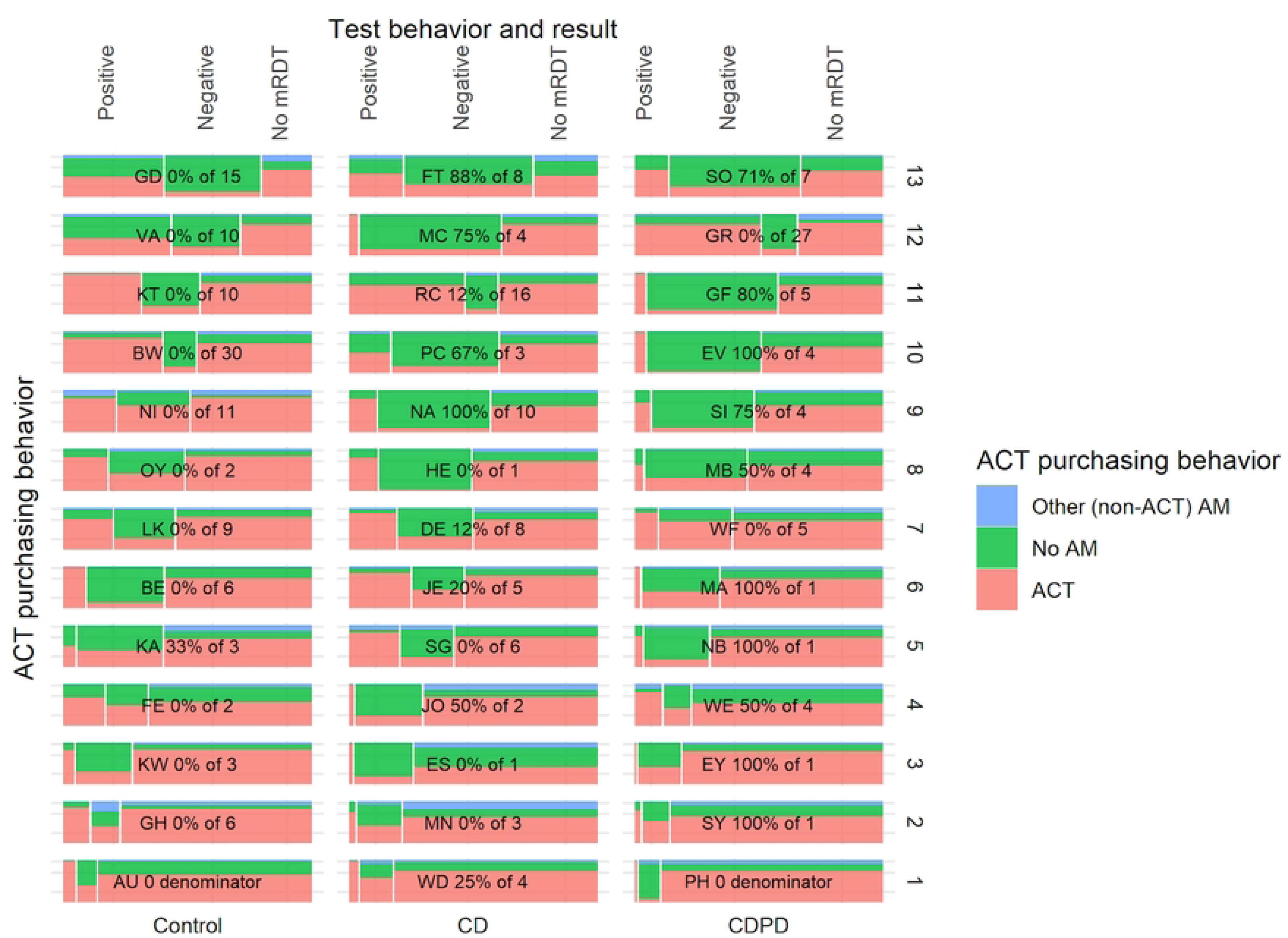
Outlet level heterogeneity in malaria testing and antimalarial drug dispensing in each arm. Outlets are grouped in columns by arm and ordered by proportion tested from highest (top) to lowest (bottom). Outlet testing rates ranged from as little as 10% to more than 75%. Likewise, ACT dispensing to test-negative patients showed stark differences between shops. Less heterogeneity is observed in ACT dispensed to untested clients.

Although integration of testing into their business practices was highly heterogenous, the use of ACT by untested clients was uniformly high with much less variation between clusters (Figure 3). Testing rates increased across all arms following client-directed sensitization via CHWs particularly in the intervention arms where it increased from 33% to 51% and 36% to 63% in the CDPD and CD arms respectively, indicating that outlets may not have been consistently offering information about mRDT testing to their clientele (Figure 4).

**Fig 4:**
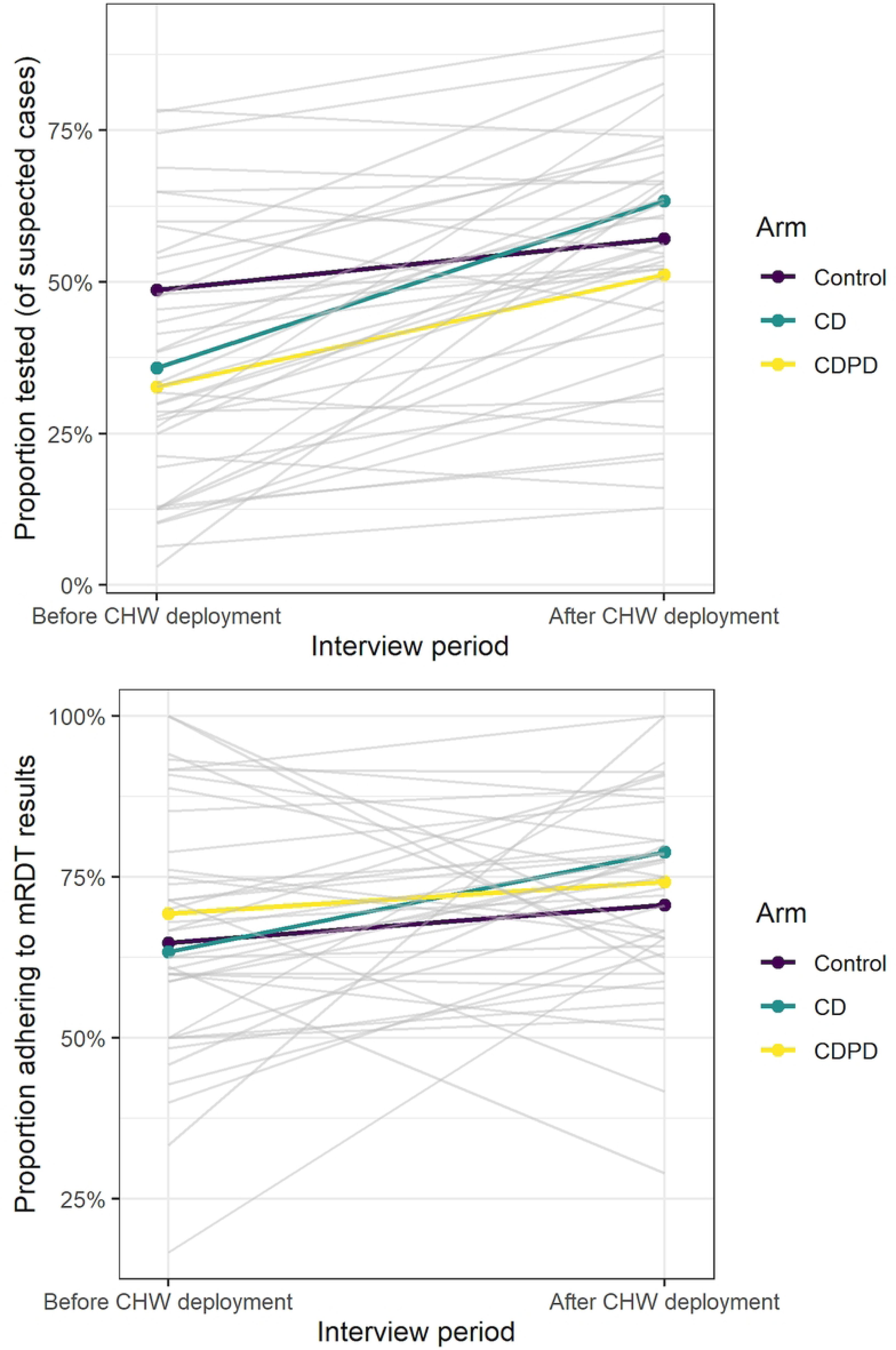
Change in outlet management of suspected malaria before and after client-facing sensitization. (a) proportion of clients tested and proportion of clients adhering to test results before and after. Each outlet is shown as a gray line with the arm-specific means shown in color. Few shops declined in testing rates after sensitization but impact on adherence was much more variable.

Outlets agreed to offer mRDT testing at a low fixed price to suspected malaria cases. Overall, there was good adherence to the study prescribed price of the mRDT (40 KES). Only 15% of participants reported being charged more than 40 KES, although 20% did not know the price they paid specifically for the test (Table 4).

**Table 4:** Expenditures by test result and arm.

| Expenditure (KES), mean (SD) | Study Arm |  |  |  |
| --- | --- | --- | --- | --- |
|  | Control | CD | CDPD | Overall |
| Total expenditure, All interviewees | 268 (265) | 251 (335) | 245 (268) | 254 (290) |
| Positive test at outlet | 423 (277) | 571 (532) | 498 (424) | 486 (410) |
| Negative test at outlet | 295 (246) | 286 (348) | 243 (215) | 273 (277) |
| Positive test from elsewhere | 348 (368) | 256 (288) | 297 (258) | 299 (293) |
| Negative test from elsewhere | 273 (308) | 147 (114) | 209 (205) | 202 (207) |
| No test | 146 (154) | 130 (101) | 157 (187) | 145 (153) |
| Positive test at outlet and free AL confirmed | 75 (7) | 157 (218) | 150 (153) | 151 (186) |
| **Expenditure reporting information, n/N (%)** |  |  |  |  |
| Unknown total expenditure, All interviewees | 147/1,692<br>(8.7%) | 170/1,789<br>(9.5%) | 214/2,214<br>(9.7%) | 531/5,695<br>(9.3%) |
| Unknown price of test, Tested at outlet | 222/815<br>(27%) | 131/801<br>(16%) | 129/811<br>(16%) | 482/2,427<br>(20%) |
| Unknown price of ACT, Positive test at outlet | 99/182<br>(54%) | 88/148<br>(59%) | 52/108<br>(48%) | 239/438<br>(55%) |
| Unknown price of ACT, Negative test at outlet | 36/94<br>(38%) | 42/112<br>(38%) | 44/121<br>(36%) | 122/327<br>(37%) |
| Unknown price of ACT, No test at outlet | 42/528<br>(8.0%) | 54/593<br>(9.1%) | 132/746<br>(18%) | 228/1,867<br>(12%) |
| Free AL, Positive test at outlet | 2/182<br>(1.1%) | 27/148<br>(18%) | 25/108<br>(23%) | 54/438<br>(12%) |

Central to the intervention was the offer of fully subsidized AL conditional on a positive mRDT. Intervention outlets agreed to implement the intervention by providing free AL to clients who had a positive mRDT result at the outlet. In order to understand the extent to which outlets adhered to these conditional subsidies, we examined the proportion of mRDT-positive clients who reported receiving free AL. Although all clients with a positive test in the intervention arms were eligible for a free AL, only 21% reported receiving a free AL (Table 4) and 75% of all free AL was dispensed in the time period *after* the client-facing sensitization. These results are somewhat inconclusive because 55% of test-positive clients did not know the price they paid for their ACT (AL or other ACT). In contrast, only 12% of untested clients and 37% of test-negative clients were unaware of the price they paid for their ACT (Table 4), suggesting some intended or unintended ambiguity in pricing specifically after a positive test.

Most clients (91%) were able to report the total amount of money spent at the visit to the outlet. Among those reporting a positive malaria diagnostic test at the outlet, the total spent inclusive of test and all drugs, was on average 75-148 KES *higher* in the intervention arms (423 in control vs 498 or 571 KES, in CDPD and CD respectively Table 4). Only clients who confirmed receiving a free AL paid less than clients in the control arm (150 KES or 1.5USD in CDPD compared to 423 KES or 4.2USD for positives in the control arm, Table 4).

Other malaria case management practices may have created competition between the intervention and other services provided by the outlet. Slightly more than 8% of clients reported receiving an injection (Table S4). The proportion receiving an injection was highest among test-positive clients (22%). Artemisinin injections or ‘malaria injections’ (presumably antimalarials) were the most commonly reported injections (19% and 18%, respectively) although more than half of clients who received an injection could not specify the drug (and no packaging was available for verification). Oral ACT was not commonly dispensed with an injection.

## Discussion

The goal of the TESTsmART trial was to improve antimalarial stewardship in the retail sector, which has the largest market share of antimalarials in sub-Saharan Africa. In our study, we demonstrate robust testing rates among clients with suspected malaria and substantially improved ACT targeting among tested clients. Approximately 50% of previously untested clients were tested in study outlets, which is consistent with other implementation studies in the retail sector where mRDTs were not free to the client.^27–30^ Clients were willing to pay for a test prior to treatment, although purchase of an mRDT at the outlet was slightly lower among the poorest wealth quintile. This could indicate that further reducing the price of the test may increase uptake, as was seen in some studies which offered free mRDTs at retail outlets.^31, 32^ However, this is far from certain given the nearly universal uptake among clients offered testing at the same price in a related pilot study.^16^

Information from the mRDT improved targeting of ACTs, particularly reducing consumption among those with a negative test. The adherence to test results observed in our study is comparable to studies from the formal health sector ^33^ adding to the evidence that case management is not compromised in retail venues compared to formal health facilities. We observed in our study that a quarter of clients who tested positive at the outlet did not receive an ACT (27%). This was particularly unexpected in intervention arms where AL treatment should have been free to the client. This points to either low confidence in AL as an effective therapy among providers and their clients or, more likely, inconsistent adherence to the subsidy program. The very high observed use of AL among untested clients (67%) also supports the latter explanation. We also noted that 22% of test-positive clients reported receiving injections which could explain why ACT consumption was lower in this group.

We did not find evidence that the client-or provider-directed incentives offered in our study were effective in improving case management in intervention outlets as compared to the outlets in the control arm. However, it is possible that the lack of effect of the conditional subsidy is attributable primarily to poor adherence to the study prescribed subsidy program. There seems to have been very little pass-through of the conditional ACT subsidy to the client based on client reporting of out-of-pocket costs. Given the favorable results from a community-based testing program that distributed a conditional ACT subsidy to the client via retail sector vouchers, we conclude that well-designed incentive programs could be effective but might require direct delivery of the conditional subsidy to the client rather than through the outlet. It is also possible that the provider-directed testing incentives were too modest to increase testing beyond that observed in the control arm, particularly if it was not enough to offset the lost sale of an ACT. Burchett et al reviewed 10 studies related to implementing RDTs and propose that RDT uptake and adherence is maximized when RDT use was ‘aligned well with providers’ own priorities’^34^. The private for-profit outlets that participated in our study are businesses and it may simply be that we did not find the right combination of incentives that resulted in favorable ‘alignment’ with outlet priorities. One aspect of poor ‘alignment’ could be the delay between dispensing an AL and reimbursement several days later. Anecdotal feedback from the outlets suggests that this delay could have led outlets to dissociate payments from dispensing behavior. The possibility that outlets pocketed reimbursements intended for clients is consistent with results from a study in Uganda that described outlet owners as surviving in a very competitive and crowded sector. The majority of outlets are likely not consistently profitable leading to ‘problem-solving corruption’^35^ where the context in which they operate makes it infeasible to follow some rules, whether it be the rules of our subsidies, rules about selling antibiotics or the restrictions on giving injections.^35^

The differences in out-of-pocket costs for clients with a test are striking and are worth examining in light of 1) discussion regarding how incentives might reduce out of pocket costs for clients in the retail sector and 2) the concern on the part of retailers that performing mRDTs could reduce revenue from sales of ACTs. Although it is certainly true that fewer clients purchase ACT after a negative test, our results show that these clients spend nearly twice as much as untested clients even when they don’t purchase an ACT. Furthermore, those with a positive test spend on average more than three times more than those without a test. It is possible that clients choose not to test if they have a limited budget that can only cover the cost of the ACT and we cannot assume that, if tested, they would spend as much as other clients who chose to be tested. Further, we do not know the profit margin for the outlet on non-AL drugs compared to the mRDT and AL. Nonetheless, we find that 20% of clients who confirmed receiving a free AL still spent more on average than an untested client. Overall, these results cast doubt on the concern about lost revenue due to testing or on the prospect of reducing out-of-pocket costs through testing. Although these observations seem like they would encourage providers to offer testing, the increase in testing rates following the client-directed sensitization suggests that testing was not being offered consistently by the outlet attendants.

A strength of our study design is the use of exit interviews of randomly selected clients on the day of their visit to the outlet. This approach allowed us to minimize bias that is likely to arise when outcomes are based on reports generated by the outlet, particularly when some outcomes are discouraged or against ‘the rules’. There are important limitations to consider both in our intended approach and in the study. First, interventions designed to bolster testing in retail outlets can only reach people who visit the outlet and will always miss a significant proportion of suspected malaria cases being treated with drugs purchased on their behalf, for example parents buying medication for a sick child. With respect to our study, we were unable to determine whether the intervention failed to lead to measurable improvement in case management because the incentives were not valuable enough (ineffective intervention) or because the intervention as it was designed was not implemented. In our previous trial, information from a diagnostic test was given directly to the client as was the voucher for AL (conditional incentive). This strategy worked well in reducing unnecessary consumption of ACTs even though the incentive level was comparable (mRDT was free but ACT price was subsidized to 40 KES compared to the current study where the mRDT cost 40 KES and the ACT was free). In addition, test results and out of pocket costs were self-reported by the client and may be inaccurate if communication with the client was not clear or was incomplete.

## Conclusion

A sizable proportion of clients visiting retail outlets are willing to test for malaria resulting in better targeting of ACTs among tested clients. Yet, providing client-directed subsidies through outlets or financial incentives to outlets may not result in additional improvements in testing and treatment if the incentives are not well understood or well aligned with outlet priorities.

## Funding

Research reported in this publication was supported by the National Institute Of Allergy And Infectious Diseases of the National Institutes of Health under Award Number R01AI141444. The content is solely the responsibility of the authors and does not necessarily represent the official views of the National Institutes of Health. The funders had no role in study design, execution or analysis of the data.

## Data Availability

Dataset will be published in https://research.repository.duke.edu/

NA

## Notes

### Competing Interest Statement

The authors have declared no competing interest.

### Clinical Trial

ClinicalTrials.gov (NCT04428307)

### Clinical Protocols

NA

### Author Declarations

The study was reviewed and approved by Duke University Institutional Review Board (Pro00104256) and Moi University Institutional Research and Ethics Committee (IREC/2019/304). The study is registered in ClinicalTrials.gov (NCT04428307).

